# Is early initiated physical rehabilitation exercise superior to no physical rehabilitation exercise following total hip arthroplasty? A systematic review and narrative synthesis

**DOI:** 10.1101/2025.01.07.24319217

**Authors:** Merete Nørgaard Madsen, Lone Ramer Mikkelsen, David Høyrup Christiansen, Helle Kvistgaard Østergaard, Thomas Bandholm

## Abstract

**Background:** Physical rehabilitation exercise (PRE) is commonly prescribed in the early stage after total hip arthroplasty (THA). However, systematic reviews investigating the effectiveness of PRE have reported diverse results, and often included trials using PRE in both groups or initiated at a later stage after surgery, which does not reflect clinical practice. Therefore, the study objective was to investigate whether early initiated PRE following THA was superior to no PRE in terms of improving function, reducing pain and increasing quality of life at end of intervention and 12 months after surgery.

**Methods:** A systematic review of randomized controlled trials (RCT) was conducted. Included studies were RCTs comparing PRE initiated within 3 months after primary THA due to osteoarthritis with no PRE. MEDLINE, Embase, Cinahl, Cochrane and Pedro were searched for published articles, while Scopus, Web of Science, Clinical Trials.gov and WHO International Clinical Trials Registry Platform were searched for conference papers and pre-registered trials. Study methodology was assessed by Cochrane Risk of Bias 2 (RoB2) tool and overall quality of evidence by the Grading of Recommendations Assessment, Development and Evaluation approach (GRADE). Incomplete outcome data and heterogeneity among studies precluded meta-analysis. Thus, data synthesis using vote counting was applied and tested by the binomial probability test. The results were narratively presented in text and tabular form distributed on pain, patient-reported function and performance-based function.

**Results:** A total of 10742 references were screened. Three trials (two published papers and one conference abstract) with a total of 151 participants were included in the narrative synthesis. Only outcomes at end of treatment (ranging from 3-18 weeks after surgery) were available. The observed direction of effect favored PRE in the single study reporting patient-reported function, in both studies reporting pain and in two out of three studies reporting performance-based function. However, the testing did not show significant evidence of effect of PRE. Overall, a high risk of bias was present, and quality of evidence was very low.

**Discussion:** Limited and very low quality of evidence showed no clear benefits or harms of PRE. Hence, no conclusions on superiority of PRE to no PRE can be drawn. High quality randomized controlled trials are needed to determine the effectiveness of using PRE early after THA.

**Other:** Registration: PROSPERO, CRD42020203574

## Introduction

### Rationale

Total hip arthroplasty (THA) is a common surgical intervention, with more than 1 million procedures performed per year (1). The most common indication for THA is advanced hip osteoarthritis, pain and impaired function (2–4) and overall, the surgery leads to patient improvements in pain, function and health-related quality of life (5–7). However, patients’ muscle strength and functional ability are nevertheless substantially reduced after surgery (7–12) and postoperative physical rehabilitation exercise (PRE) is commonly initiated early after surgery as recommended in clinical guidelines (13, 14).

Although PRE is recommended (13, 14), the evidence for its effectiveness is inconclusive. In a systematic review with meta-analyses by Saueressig et al., rehabilitation exercise compared to usual care, or no or minimal intervention did not significantly improve patient-reported function or hip muscle strength (15). This was the case at four specified time-points distributed from 4 weeks to 1 year after surgery (15). In contrast, other systematic reviews with meta-analyses have reported that rehabilitation exercise may be superior to standard care or no treatment, when measured on gait variables (16–19), hip abductor strength (18, 19), self-reported function (16, 18) and pain reduction (16, 18). Among the mentioned meta-analyses showing a beneficial effect of PRE, only Tang et al. presented their results within specified time-frames (16). In the short-term follow-up period (<6 months) their results significantly favored the intervention group, while the improvements were not significant at long-term follow-up (six to 12 months) (16). Besides the systematic reviews referenced above reaching different conclusions, the heterogeneity of trials included in the reviews adds to the uncertainty of the evidence for effectiveness of early initiated PRE. The trials differ in comparators (e.g. pooling usual care, minimal or no rehabilitation) with the majority of trials using PRE in their control groups, time point initiated after surgery and whether or not the participants had performed exercises between discharge and initiation of the trial. Hence, evidence specifically reflecting the fundamental clinical effectiveness of early initiated PRE (i.e. PRE being superior to no PRE) remains unclear. Therefore, there is a need for a systematic review that assesses the effect of early initiated PRE following THA, reflecting the time point used in clinical practice and with a comparator with no PRE. Hence, our research question and objective are as follows.

### Objective

To investigate whether early initiated PRE is superior to no PRE following THA in terms of patient-reported outcomes for function, pain and quality of life or performance-based outcomes for function, when measured at end of intervention and 12 months after surgery.

### Research question

Is early initiated physical rehabilitation exercise superior to no physical rehabilitation exercise following total hip arthroplasty in terms of patient-reported outcomes for function, pain and quality or performance-based outcomes for function?

### Definitions

The definition of PRE is inspired by and closely related to PubMed’s MESH term “Exercise Therapy” (20).

#### PRE

Physical activity and/or exercise, that is designed and prescribed by a health professional to be performed after discharge, for restoring function or reducing pain caused by disease, injury or surgery.

#### no PRE

None of the above but may include encouragement to stay active, continue life as usual and resuming activities when feeling ready.

#### Early initiated

Within three months after index surgery.

#### Immediately initiated

Within one month after index surgery.

## Methods

### Design

A systematic review following the guidelines in Cochrane Handbook for Systematic Reviews of Interventions (21) was performed. The study protocol was based on the PRISMA-P guidelines (22) and the study was pre-registered in PROSPERO (The International prospective register of systematic reviews) (23). Study reporting adheres to the Preferred Reporting Items for Systematic Reviews and Meta-analyses (PRISMA) statement (24) and The synthesis Without Meta-analysis (SWiM) reporting guideline (25).

### Eligibility criteria

Criteria for study inclusion were defined based on population, intervention, comparison, outcomes and type of study (PICOT) as shown in Table 1. No limitations in publication year or language were applied.

**Table 1.** Eligibility criteria.

|  | Inclusion criteria | Exclusion criteria |
| --- | --- | --- |
| **Population** | Patients following unilateral THA due to hip osteoarthritis | Age below 18 years |
| **Intervention** | PRE was the main intervention. Other supplemental interventions (such as electric stimulation, acupuncture, or continuous passive motion) were only allowed as an adjunct to PRE | - PRE was commenced later than three months after index surgery.<br>- Some kind of PRE had already been performed between discharge and study start. |
| **Comparison** | No PRE | Prescription of another modality (e.g. electrical stimulation) aiming to increase function and reduce pain. |
| **Outcomes** | At least one of the following outcome domains were reported:<br>Pain (hip-related)<br>Patient-reported function<br>Performance-based function<br>Health related Quality of Life<br>Adverse events | Outcomes were measured outside the eligible time frames, which were:<br>- end of treatment (+/- 2 weeks)<br>- 12 months after surgery (+/- 6 months)* |
| **Type of studies** | Randomized controlled trials |  |

THA: Total Hip Arthroplasty; PRE: Postoperative Rehabilitation Exercise.
\* If outcome was measured at several time points, data from the follow-up closest to 12 months was used

### Search strategy

Systematic searches were undertaken in October 2020 and updated between January 17 2024 and January 31 2024. The databases MEDLINE, Embase, CINAHL, PEDro and Cochrane Central Register of Controlled trials (CENTRAL) were searched for published papers, while Scopus and Web of Science were searched for conference papers. Pre-registered trials were searched using ClinicalTrials.gov and the WHO International Clinical Trials Registry Platform portal.

The search string was developed based on the eligibility criteria for population, intervention and type of study. Further details on the search strategy and an example of a specific search history (the updated MEDLINE search) is presented in Appendix A. The search strategy was validated by checking that expected relevant studies identified through recent (within the last 10 years) systematic reviews were identified. Additionally, reference lists were checked, and citation searches of the included studies were performed in Web of Science. No further studies were identified during this process.

### Study selection

The results were downloaded to EndNote (Version 21, Clarivate), in which duplicates were automatically identified and removed after being manually checked. The remaining studies were uploaded to Covidence (26), where further duplicates were automatically identified and removed after manual check. The Covidence software was used in the remaining selecting process.

Using the eligibility criteria presented in Table 1, two reviewers (MNM and HKØ) independently screened the studies based on title and abstract. If at least one of them considered a trial to potentially be eligible, it was evaluated in full text by the same two review authors. They independently assessed the eligibility criteria in order of importance, and the first “no“-response was used and registered as reason for exclusion (27). In this review, the pre-defined order of importance was as follows: a) study type, b) population, c) intervention, d) comparison, e) outcome.

Consensus between the two review authors was reached by discussion and when in doubt by involving a third reviewer (TB). In case of insufficient information to judge the eligibility of a study, the corresponding author of the study was contacted for clarification. In case of papers being reported in foreign languages not familiar to the authors, eligibility was decided by assistance from bilingual clinicians and researchers contacted at either Silkeborg Regional Hospital or using Cochrane TaskExchange (now Cochrane Engage) (28). In three cases, it was not possible to decide with certainty whether the studies fulfilled the eligibility criteria. Those studies were neither included nor excluded, but were presented separately in the result section in the table “Characteristics of studies awaiting classification” (27).

### Data collection and outcomes

MNM and HØ independently extracted trial data (authors, country and year of publication, trial identifier and registry, study design, number of participants, duration of study, funding), population-related data (gender, age, comorbidities, self-efficacy, preoperative activity level, preoperative pain and preoperative function), and outcome data (baseline and follow up measures of function, pain and quality of life). Furthermore, The Consensus on Exercise Reporting Template (CERT) (29, 30) was used to extract and report exercise interventions from the included trials (29). Consensus was reached by discussion. Data within each outcome domain were sought for and extracted at end of intervention and/or at follow-up closest to 12 months as stated in Table 1.

In all included studies, some relevant data were missing, incomplete or inconsistent (31–33). The corresponding authors were therefore contacted but besides obtaining sample sizes from one study (32) it did not lead to clarification. Instead, where possible, available data were used to calculate between-group differences as described in the following. In one study, two relevant intervention groups were identified, and in each group, the post-intervention outcomes were reported in further two groups (32). Similarly, the control group’s outcome was reported split in two (32). As recommended by the Cochrane Network, we therefore pooled data for the four estimates from the intervention groups and thereafter the two estimates for the control group to be able to calculate mean difference between the pooled intervention and pooled control group (34). The estimates in each group were weighted according to the obtained sample size from the author. Additionally, in another study, we estimated mean difference between groups based on reported post-intervention estimates (31). Since both studies presented their post-intervention estimates with mean and standard deviation (SD)(31, 32), we assumed a parametric distribution of data and used two-sided t-test to estimate mean difference between groups with corresponding 95% confidence intervals and p-values.

#### Primary outcomes (critical)

The primary outcomes were patient-reported outcomes for function and pain measured at the end of intervention. Function and pain were chosen, due to a) the definition of PRE, where the two outcome constructs were the main reasons for prescribing the intervention and b) the overall reason for THA surgery is to improve function and reduce pain (2). Patient-reported instead of performance-based measuring of function were chosen, because patient-reported outcome measures (PROMs) are considered the best tools available to measure patient-centered outcomes objectively (35). This in accordance with The World Health Organization (WHO), which has defined patient-centeredness as a fundamental characteristic for the quality of healthcare (36).

#### Secondary outcomes (important)

Secondary outcome measurements included patient-reported outcomes for function and pain 12 months after surgery as well as performance-based measure of function, health related quality of life and adverse events – measured at either end of intervention or 12 months after surgery. Follow-up measures of adverse events with or without causality to the intervention were searched for. This included search for available adverse event definitions, how information was collected, number/proportion (absolute risk) of cases per arm and number/proportion of withdrawals in each group due to adverse events (37).

An inclusive strategy was used, thus all types of outcome measurements in each domain could be included. If more estimates were reported for the same domain, only the one considered most relevant was included in the synthesis. The pre-defined hierarchy of relevance for frequently used outcome measurement tools is presented in Table 2. In one study (32), several performance-based function measurements were reported, but none were among the pre-defined tools. To decide which was the most relevant, two of the authors (TB and LRM) independently and blinded to the results judged this and came to the same result. Hence, data was extracted on the variable E_area_ in unipedal stance, which is a measure of postural function, with higher value (area) indicating lesser stability (32).

**Table 2.** Pre-defined hierarchy of frequently used outcome measurement tools.

| Outcome domain | Outcome measurements – presented in order of relevance | Comments |
| --- | --- | --- |
| **Patient-reported function** | HOOS (subscale ADL), WOMAC (physical function), SF-36 (physical function or PCS), SF-12 (PCS), OHS, HHS | Specific subscales of function have higher priority than overall scores |
| **Pain (hip-related)** | HOOS (pain), WOMAC (pain), VAS, NRS, SF-36 (pain), OHS, HHS (pain item) | If there is no description on pain location, it is assumed to be hip-related |
| **Performance-based function** | Walk test, chair stand test, stair climb test. Isometric hip muscle strength (abduction, flexion, extension, adduction) | Core tests recommended by OARSI have higher priority than muscle strength |
| **Health-related quality of life** | HOOS (QOL), OAKHQOL, SF-36, EQ-5D | Disease-specific outcomes higher than generic |

Abbreviations: HOOS: Hip injury and Osteoarthritis Outcome Score; ADL: Activities of Daily Living; WOMAC: The McMaster Universities Osteoarthritis Index; SF-36: 36-item Short Form Survey; PCS: The Physical Component Summary; SF-12: 12-item Short Form Survey; OHS: Oxford Hip Score; HHS: Harris Hip Score; VAS: Visual Analogue Scale; NRS: numeric rating scale; 6MWT: 6 minute walk test; OARSI: Osteoarthritis Research Society International; OAKHQOL: The osteoarthritis knee and hip quality of life questionnaire; EQ-5D: European Quality of Life – 5 Dimensions.

### Risk of bias

Risk of bias for each result (outcome level) was assessed using the Revised Cochrane risk-of-bias tool for randomized trials (RoB2) (38–40). Assessment included bias in the following domains: randomization process, deviations from intended interventions, missing outcome data, measurement of the outcome and selection of the reported result. Using the algorithm included in RoB2 (40), each of the mentioned domains were evaluated and assigned to one of three levels: low risk of bias, some concern or high risk of bias (38–40). Subsequently an overall risk of bias judgement for the result, using the same three levels was reached (38–40). Two review authors (MNM and HKØ) independently evaluated the risk of bias domains and disagreement between the reviewers was solved by consensus or by involving a third reviewer (TB).

### Synthesis methods and certainty assessment

A narrative summary of evidence is presented in text along with a table presenting the results from each study in each of the outcome domains. Hence, the grouping of studies is kept as originally planned if meta-analysis could have been undertaken. Due to incomplete outcome data and inconsistent effect measures across studies, direction of effect was used as standardized metric and vote counting as synthesis method (41). Vote counting is purely based on direction of effect and does not take into account effect size, statistical significance or sample size. The method only answers if there is any evidence of effect, which was tested using two-sided binomial probability test (41). In the table, the observed direction of each effect estimate is presented along with our summarized direction of effect and quality of evidence within each outcome domain. This domain specific direction of effect was primarily based on the binomial probability test and supplemented by evaluating the magnitude of effect size and statistical significance of the included results. Higher weight was given to results from studies being published in peer-reviewed papers, being pre-registered, using recommended outcome measurements or having higher generalizability by using common interventions. The overall quality of evidence of each of the outcome domains was evaluated according to Grading of Recommendations Assessment, Development and Evaluation (GRADE), where domains such as risk of bias, inconsistency, indirectness, imprecision and publication bias graded the overall evidence as high, moderate, low or very low (42–46).

### Deviations from protocol

A meta-analysis was planned, but due to incomplete outcome data and concerns of clinical and methodological heterogeneity between studies (based on comparison of study characteristics, Table 3), this was not considered reasonable (41). Therefore, in accordance with our pre-registered plan (PROSPERO), a narrative summary of evidence is instead presented in text and tabular form. The choice of vote counting as synthesis method was not pre-defined but based on data availability. Also, due to a small number of studies eligible for inclusion, the sub-analysis of subgroups was precluded. Finally, due to the thorough search and small number of studies included, no further reporting bias assessment was performed.

**Table 3.** Trial characteristics and results of the included randomized controlled trials.

| **Publication status** | **Trial**<br>Author, Year<br>Country<br>Study design<br>Trial registration and identifier | **Participants (intervention/ control)**<br>Numbers<br>Age (years)<br>Gender<br>Other* | **Intervention**<br>Initiating point<br>Duration<br>Short description† | **Control**<br>Short description | **Outcomes**<br>Reported outcomes distributed on applicable domains‡ | **Follow-up**<br>Time points | **Main results (as reported by authors)** |
| --- | --- | --- | --- | --- | --- | --- | --- |
| Published paper | Monaghan et al. 2017 (31)<br>Ireland<br>Parallel RCT<br>Clinical trial.gov<br>NCT01683201 | N= 32/31<br>Age in mean (SD)<br>68 (8)/69 (9)<br>Gender (proportion of females):<br>37%/26% | Initiated 12 weeks after surgery<br>Duration: 6 weeks<br>Physiotherapist supervised exercise classes 35 minutes twice a week. The exercise programme consisted of 12 functional exercises which were progressed as necessary | Usual care - no postoperative physical rehabilitation exercise | *Patient-reported function*<br>WOMAC function<br>*Pain*<br>WOMAC pain<br>*Performance-based function*<br>6MWT | 18 weeks after surgery (end of intervention) | At 18 weeks: Mean difference with 95% CI<br>WOMAC function: -4.0 (-0.71 to 1.0)<br>WOMAC pain: -0.81 (-1.8 to 0.2)<br>6MWT: 21.9 (0.60 to 43.3) |
| Published paper | Martinez et al. 2022 (32)<br>France<br>Parallel four-arm RCT<br>No registration identified | Intervention group estimates are based on pooled data from two groups<br>N=27/14<br>Age in mean (SD)<br>68.9 (9.4)/<br>69.6 (5.8)<br>Gender (proportion of | Initiated 10-21 days after surgery<br>Duration: 3 weeks<br>Two intervention groups<br>Rehabilitation on Stabilometric Platform (SP) group (RSPG): Supervised exercise program using an SP. Three sessions per week, comprising 13-14 exercises, each with a duration between 10 and 120 seconds (overall | No rehabilitation control group (THACG)<br>Walk as often as possible. No further instructions. | *Performance-based function*<br>E<sub>area</sub> – unipedal stance | 45-60 days after surgery (end of intervention) | Mean (SD) E<sub>area</sub> – unipedal stance at follow-up.<br>RSPG:<br>Preferred operated leg: 6.44 (3.39)<br>Nonpreferred operated leg: 11.02 (10.14)<br>SDHRG:<br>Preferred operated leg: 10.45 (3.58)<br>Nonpreferred operated leg: 7.14 (0.06) |
|  |  | females):<br>56%/50% | duration of 15-17 minutes)<br>Self-directed home-based rehabilitation group (SDHRG): Self-directed home-based exercises after an initial instruction. Exercises 6 days per week. Recommended 20-30 minutes of walk per day. |  |  |  | THACG:<br>Preferred operated leg: 6.48 (3.38)<br>Nonpreferred operated leg: 7.74 (1.23) |
| Conference abstract | Marcu et al. 2019 (33)<br>Romania<br>Parallel RCT<br>No pre-registration identified | N=24/23 | Initiated 7-10 days after surgery<br>Daily physical exercise<br>Duration: 2 weeks | Analgesics and NSAID's when needed | *Performance-based function*<br>Muscular strength (no further description)<br>*Pain*<br>VAS (100 mm) | 3-4 weeks after surgery (end of intervention) | *Muscular strength*<br>Intervention group improved 8.3%<br>Control group did not improve<br>Difference between groups: $p < 0.05$<br>*Pain*<br>Intervention group improved 42.6%<br>Control group improved 33.4%<br>Difference between groups: $p = 0.000054$ |
| Ongoing | Mark-Christensen 2021 (47)<br>Denmark<br>Parallel three-arm RCT<br>NCT04960241 | No information (A total number of 168 is planned for) | 5 -7 days after surgery<br>Two intervention groups:<br>Home-based telerehabilitation: Interactive virtual rehabilitation using a mobile app. | No physical rehabilitation. Will receive encouragement to stay active and to gradually return to activities of daily living | *Patient-reported function*<br>HOOS ADL<br>*Pain*<br>HOOS pain<br>*Performance-based function*<br>40m walk test | 6 weeks(end of intervention), 3 and 12 months after surgery | No results |
|  |  |  | Home-based rehabilitation: Exercises illustrated in a written exercise-program performed at home<br>The same four strengthening exercises will be performed in both intervention groups<br>Duration: 6 week<br>Once a day with an intensity of 15 RM<br>Progression: every second week |  | *Quality of life*<br>HOOS quality of life |  |  |

\* If data were available, comorbidities, self-efficacy, preoperative activity level, preoperative pain and preoperative function was reported
† Further description is available in appendix B, Table B.
‡ Outcome domain relevant to this review (patient-reported function, pain, performance-based function, health-related quality of life or adverse events)
Abbreviations: CI: Confidence interval; SD: Standard deviation; RCT: Randomized controlled trial; WOMAC: The McMaster Universities Osteoarthritis Index; SF-12: Short Form 12 physical health score; VAS: Visual Analogue Scale; 6MWT: 6-minute walk test; HOOS: Hip injury and Osteoarthritis Outcome Score; ADL: Activities of Daily Living;

## Results

### Study selection

After initial removal of duplicates, 10742 studies were screened for eligibility and four fulfilled the criteria (31–33, 47). Two were published papers (31, 32), one was a conference abstract (33) and one was an ongoing replication trial (47) adhering to its first trial’s protocol paper (48). Hence, three studies (based on a total of 151 participants) reported results and were included in the data synthesis.

In three cases, eligibility could not be decided with certainty (49–51), hence these studies are presented as “awaiting classification”. The selection process is illustrated in Figure 1.

**Figure 1.**
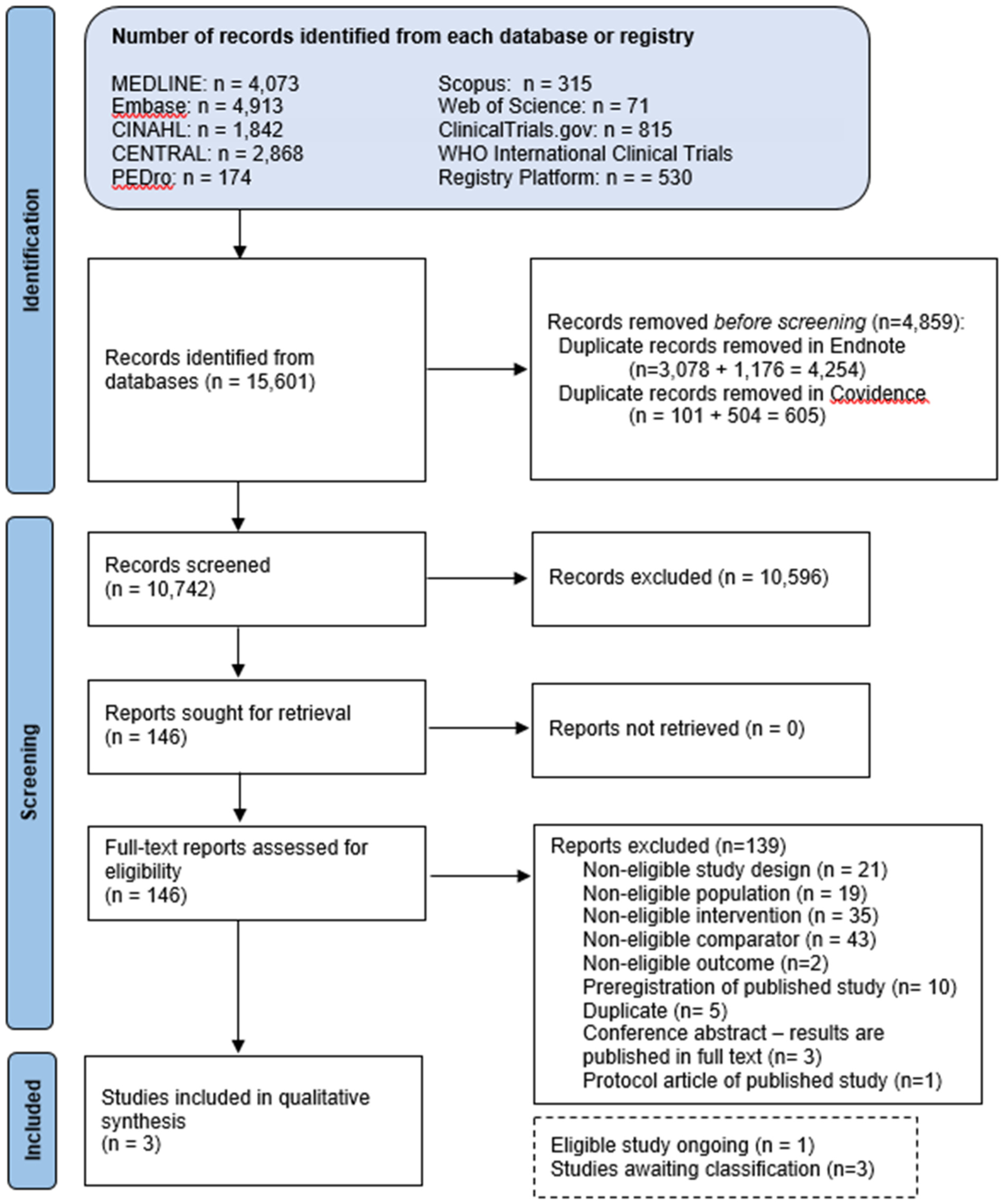
PRISMA flowchart of identification and selection of studies

Based on tables of study characteristics in previous systematic reviews (15, 16, 18, 19, 52), further studies might appear to meet our eligibility criteria (53–58). Therefore, reasons for excluding these trials are specified in Appendix B, Table A.

### Study characteristics

Two of three studies reported age and gender. Mean age in the groups were between 68 and 70 years and the proportion of females varied from 26% to 56%. The interventions were initiated between one and 12 weeks after surgery, had a duration of two to six weeks, and follow-up between three and 18 weeks after surgery. In all studies, only end-of-treatment follow-up was reported. Across studies, different outcomes measures were used. No results within the domain health-related quality of life were identified. Characteristics on each study is provided in Table 3 with intervention specific details according to CERT in Appendix B, Table B. Also, characteristics on studies awaiting classification are available in Appendix B, Table C.

## Results

In the following, study results at end-of-treatment within each outcome domain is described. Risk of bias assessment of each outcome in each study is provided in Table 4 and a summary of findings in Table 5.

**Table 4.**
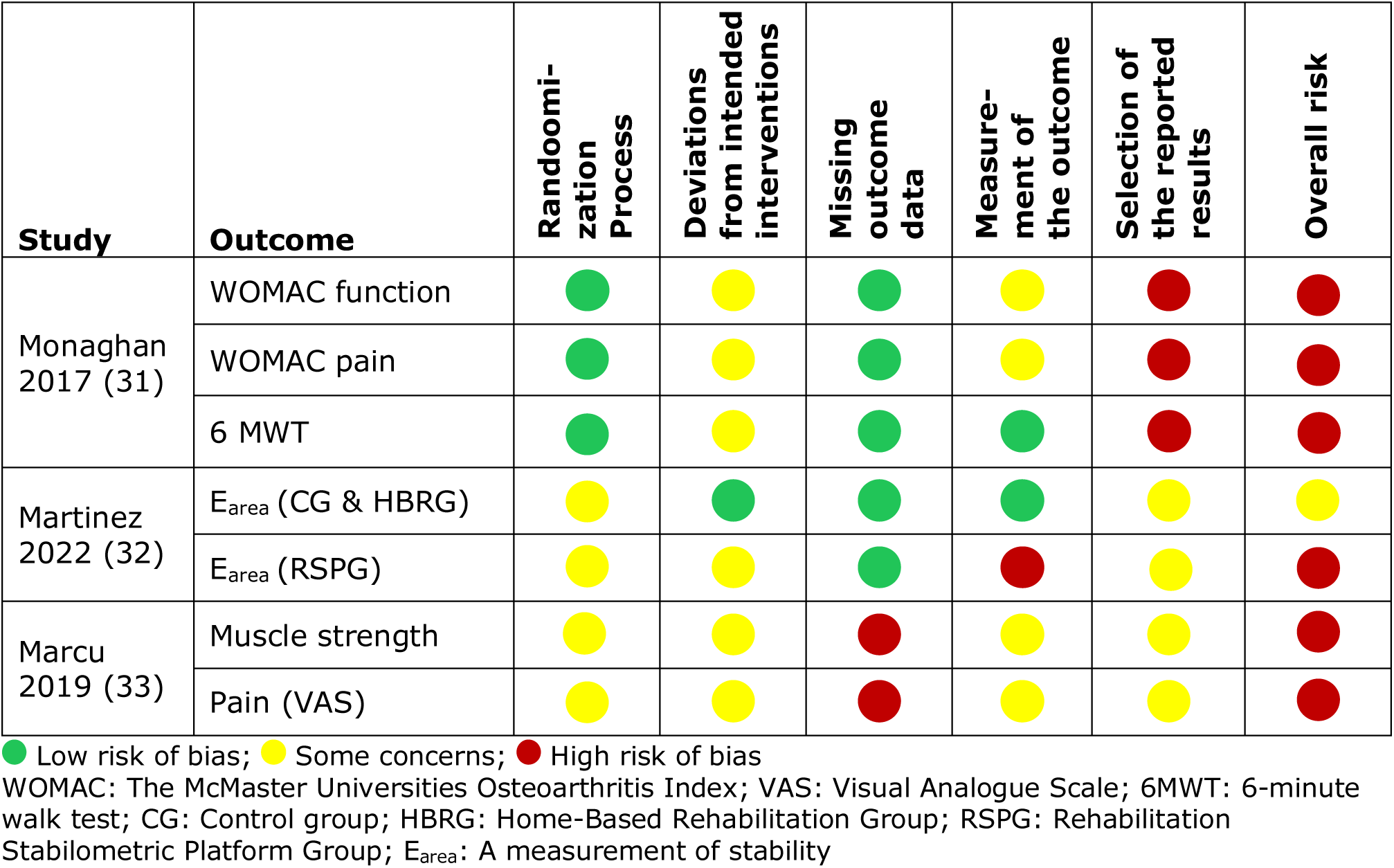
Risk of bias judgements for each outcome of the study according to the Revised Cochrane risk-of-bias tool for randomized trials (RoB2)

**Table 5.**
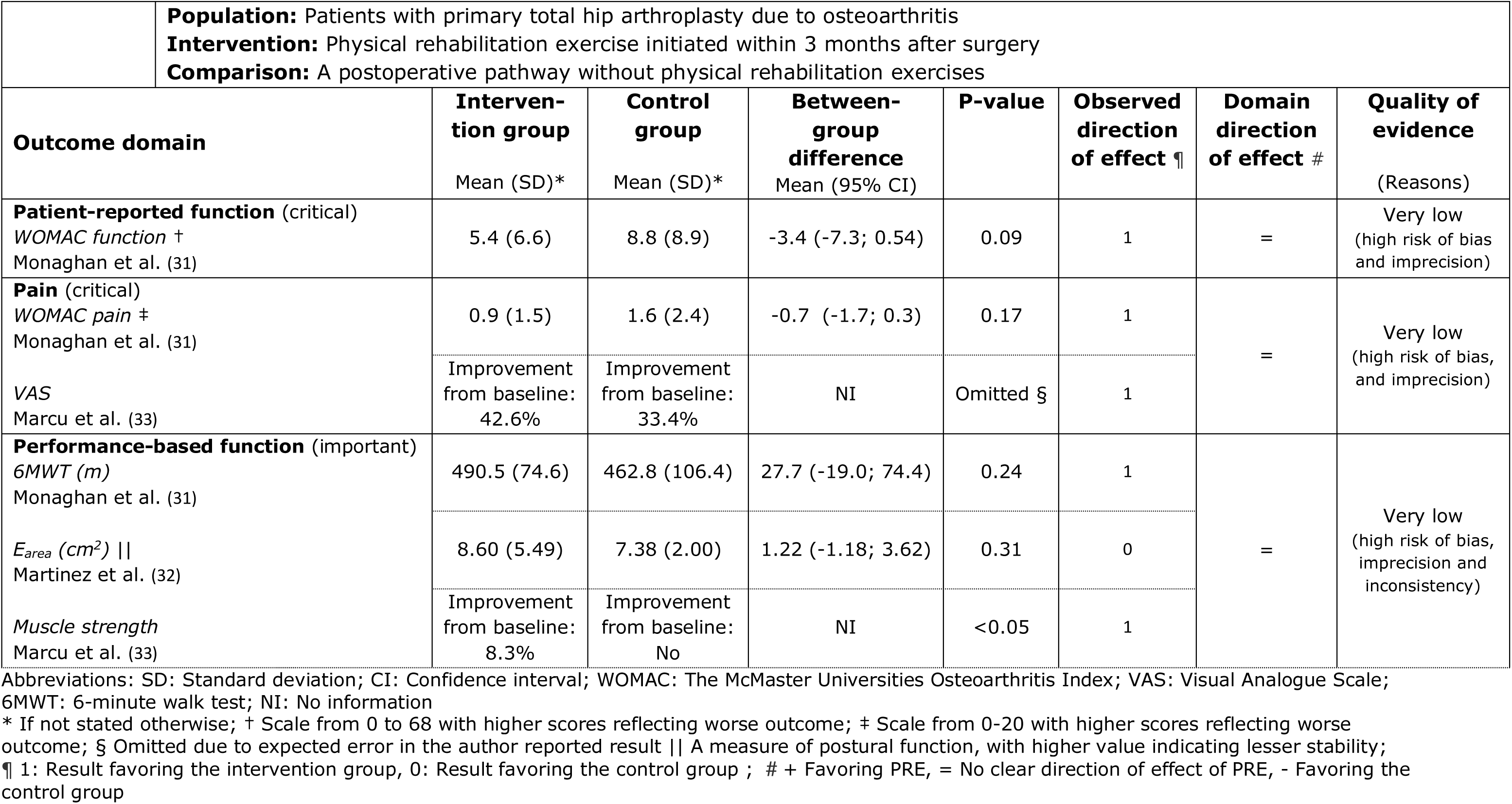
Summary of the effects of physical rehabilitation exercise at the end of treatment. In each outcome domain, separate estimates are presented for each study, while direction of effect and quality of evidence is summarized for each domain.

Due to incomplete reporting of outcome data in the conference abstract (33), in the narrative syntheses on pain and performance-based function below, we address if our overall summary of direction of effect for the domains would have differed if this study had not been included.

### Patient-reported function

Only the study by Monaghan et al. assessed a patient-reported outcome (31) using the WOMAC function subscale. When using our calculated estimate to compare their groups’ results at 18 weeks after surgery, we did not find any eligible evidence of a clear between-group difference, although the observed direction of effect favored the intervention group by having a lower (better) mean score compared to the control group (Table 6).

As only one study was included, the proportion favoring exercise was 100% (binomial exact 97.5 one-sided exact CI 2.5% to 100%, p = 1.00). Together with a non-significant between-group difference and a small effect size, our overall interpretation is that no clear direction of the effect of PRE on patient-reported function could be determined. High risk of bias was present for the outcome (Table 5).

### Pain

The paper from Monaghan et al. (31) and the conference abstract from Marcu et al. (33) reported pain level at end of intervention but differed in time-points for study start after surgery, duration of intervention, time-point for end-of-intervention and measurement tools used (Table 4a). Monaghan et al. used the WOMAC pain subscale (31) while Marcu et al. used a 100 mm VAS (33). When using our calculated estimate to compare Monaghan’s results in the intervention group and control group at 18 weeks after surgery, we did not find eligible evidence of a between-group difference but the observed direction of effect favored the intervention group. Similarly, at three to four weeks’ follow-up, Marcu et al. reported improvements from baseline in both groups and stated a p-value of 0.000054 (Table 4) which presumably represents the statistical difference between groups favoring the intervention group (Table 6) (33). No baseline or follow-up estimates nor estimates of differences between groups were available in the abstract.

High risk of bias was present for both outcomes, but especially the incomplete data from Marcu et al. (33) leads to reduced trustworthiness of the result, and we are concerned that the reported p-value of 0.000054 might be a typing error. We did not succeed in getting in touch with the authors, thus validity of the reported p-value could not be clarified by the reporting source. However, after consulting a statistician, we have reason to believe the between-group difference is non-significant.

Both studies reported results favoring exercise (100% (binomial exact 97.5% one-sided CI: 16% to 100%), p=0.50) but we did not find evidence of a clear direction of the effect of PRE on pain. This summarization would be the same if the conference abstract had not been included.

### Performance-based function

All three studies reported results concerning performance-based function, but three different outcome variables and measurement tools were used. Only Monaghan et al. (31) used one of the Osteoarthritis Research Society Internationals (OARSI) recommended performance-based tests to assess physical function in people diagnosed with hip or knee osteoarthritis (59, 60), namely the 6-meter walk test. At 18 week follow up, and after 6 weeks of exercises, the intervention group had a longer (better) post-intervention gait distance compared to the control group, but when using our calculated estimate to compare the groups, we did not find any eligible evidence for a between-group difference (Table 6) (31). Martinez et al. measured postural control at 45-60 days after surgery following three weeks of PRE (32). A stabilometric platform, which was also used as exercise tool in one of the intervention groups, was used in the assessment. At end of intervention, E_area_ was slightly lower (better) in the control group than in the intervention group, but when comparing the group results, we did not find any eligible evidence of a between-group difference (Table 6) (32). At three to four weeks after surgery, and after two weeks of daily exercises, Marcu et al. (33) reported their intervention group to have improved in muscular strength compared to no improvement in the control group. Thus, the observed direction of effect favors the intervention group (Table 6) (33). No specifications were reported on how and which muscles were tested. Additionally, no baseline, follow-up estimates or estimates of differences between groups were available in the abstract, but the authors reported a p-value <0.05 (33). Across study outcomes in this domain, a high risk of bias was present.

In summary, 2 of 3 studies favored exercise (67% (binomial exact 95% CI: 9% to 99%), p=1.00), and we did not find evidence of a clear direction of the effect of PRE on performance-based function. This summarization would be the same if the conference abstract had not been included.

## Adverse events

None of the three studies defined or addressed adverse events. However, in the study by Martinez et al. (32), one participant did not complete the intervention due to pain and were excluded from the analysis. No withdrawals or exclusions due to complications were described in the study of Monaghan et al. (31), , while Marcu et al. (33) did not provide information on this subject.

## Discussion

### Main results

Our systematic search initially performed in October 2020 was updated in January 2024, but still, only limited and very low quality of evidence was available. Synthesis using vote counting and narrative summarization did not indicate clear benefits or harms of using PRE in the early stage after THA, neither in terms of patient-reported function, performance-based function or pain measured at end of intervention.

### Comparing with previous research

To our knowledge, no other systematic reviews have reported results directly comparable to ours. Looking at other systematic reviews with meta-analysis addressing similar aspects, we identified two studies (15, 16) explicitly reporting results within the first 6 months after surgery, thus being somewhat comparable to end of intervention time-points in our studies Our results are in concordance with the meta-analysis of Saueressig et al., who investigated effect of rehabilitation exercise compared to usual care, or no or minimal intervention (15). Using results from four studies at the time-point closest to 4 weeks after surgery, they found a standardized mean difference (SMD) in self-reported function of 0.01 (-0.18 to 0.20) and hip abduction strength of -0.49 (-2.61 to 1.64) (15). Based on three studies, the corresponding estimates closest to 12 weeks after surgery were -0.08 (-0.23 to 0.07) and -0.26 (-1.28 to 0.76) (15). Hence, no eligible evidence of a between-group difference was found. It should be noted though, that comparison to our study is limited, as all studies contributing to the meta-analyses by Saueressig et al. had control groups, who performed some type of PRE according to our definition (15). In a more recent meta-analysis by Tang et al., a higher number of (but not newer) studies were included (16). At short-term follow-up (<6 months after surgery), results from 8 studies showed an SMD on self-reported function of 0.37 (0.14 to 0.60) favoring the intervention group, while the corresponding result on pain based on seven studies was 0.43 (0.17 to 0.69) (16). Standardized mean differences on functional performance and muscular strength tended to favor the experimental group but were not significant (16). Reasons for the disparity between the results from Saueressig et al.(15) and Tang et al.(16) are unclear, but may be due to using different eligibility criteria. Still, compared to our study, their results did not address the fundamental clinical effectiveness, since all trials included in their short-term follow-up meta-analyses either used PRE in the control group or had performed PRE before study start.

### Methodological considerations

We used restrictive eligibility criteria to be rather specific and direct in addressing fundamental clinical effectiveness of PRE. If we had broadened the criteria and included trials with a few weeks of PRE prior to study start, the number of eligible studies may have increased slightly, but our results would have less accuracy and generalizability to clinical practice, which is relevant if used to inform decisions on the common clinical practice of prescribing exercises directly after discharge. If we had used even more narrow criteria (e.g. only included trials initiated within a month, trial interventions using commonly used exercises and recommended outcome measures only), it would have further enhanced generalizability to clinical practice but in this case have led to no studies eligible for inclusion.

In the studies by Monaghan et al. and Martinez et. al, we calculated between-group differences based on post-intervention estimates (31, 32). Additionally, in the study by Martinez et al., we pooled data from several groups assuming equal population distribution (32). These decisions could induce uncertainties of the estimates, and in the case of Monaghan et al., it changed the statistical significance of WOMAC function and 6MWT. Due to our choice of synthesis method, it did not influence our overall interpretation of the effect of PRE, since the direction of effect was the same.

In the synthesis we used vote counting, but for the domain performance-based function, it had been possible to instead combine p-values. We tested the impact of our choice, and applying the method of combining p-values would not have led to a different interpretation of the effect of PRE in this domain. Also, if the abstract (33) had been excluded, we could instead have summarized effect estimates, resulting in a more informative synthesis. However, to be able to include all study results in the synthesis and to apply the same method across outcome domains, we consider vote counting as the best choice. It could be discussed whether it was reasonable to apply a binomial probability test on study samples of one to three studies, but we chose to do so to adhere to exemplified approach described by McKenzie and Brennan (41) and to form the basis for future updates of this review.

### Strengths and limitations

The most important limitation of our systematic review is the low number of included studies, their heterogeneity and the very low quality of evidence on each outcome domain, which precluded meta-analysis and assessment of publication bias. However, our comprehensive and highly sensitive search did not indicate the presence of publication bias but rather a lack of evidence. This underpins an existing research gap in the field, and to provide up-to-date evidence summaries, we therefore plan to regularly update the literature search and data synthesis as new evidence becomes available. Hence, our study has the character of a living systematic review (61).

Using vote counting in the synthesis is also considered a limitation. This method can only answer if there is any evidence of effect, but is of limited use, since it does not provide information on the magnitude of effects or account for relative sizes of the studies. Still, since data did not allow us to perform more robust and informative statistical synthesis when including all studies in the analysis, vote counting is considered better and more acceptable than a narrative summary based on subjective decisions alone (41).

Strengths of our study comprise pre-registration of our protocol to increase transparency, and the highly sensitive search with no restrictions in language, publication time or publication type. Furthermore, during the entire process of planning, conducting and reporting our study, we have followed acknowledged guidelines (see design section).

### Implications

Our study adds to the inconclusive evidence on the effectiveness of PRE, and especially, the small number of eligible studies reveals a gap in evidence addressing the fundamental clinical effectiveness (whether PRE is superior to no PRE), which is essential to inform future clinical practice. High-quality randomized controlled trials are needed to determine the effectiveness of using PRE early after THA.

## Conclusions

Limited and very low-quality evidence showed no clear benefits or harms of initiating PRE in the early stage after THA. Due to a small number of heterogeneous trials, high risk of study bias, imprecision and inconsistency, no conclusions on superiority of PRE to no PRE could be drawn. Further high quality randomized controlled trials are needed to determine the effectiveness of using PRE early after THA.

## Other information

### Registration and protocol

The study protocol was pre-registered in the International prospective register of systematic reviews (PROSPERO). Registration number: CRD42020203574 (23)

### Support

Sponsor: Elective Surgery Centre, Silkeborg Regional Hospital (Project ref. 654893)

Funding is received from: The Association of Danish Physiotherapists’ Fund; Graduate School of Health, Aarhus University; Department of Clinical Medicine, Aarhus University; The Danish Rheumatism Association; The Regional Hospital Central Jutland’s Research Fund and The Frimodt-Heineke Fund.

### Data availability

All data used in this study is either available in the original studies (references provided) or is presented in this paper or its supplementary material.

## Acknowledgements

The authors wish to express their gratitude to these people for their assistance in determining eligibility of studies reported in languages other than English: Yuan Chi; Zhaolun Cai, MD, PhD; Gang Chen, MD, PhD and Søren Søndergaard, MD, PhD.

