## Appendix A for "Is early initiated physical rehabilitation exercise superior to no physical rehabilitation exercise following total hip arthroplasty? A systematic review and narrative synthesis"

### Supplemental details on search strategy

The databases (MEDLINE, Cinahl, Embase, C and Pedro) were searched using keywords and text words in two blocks: 1) total hip arthroplasty (population) and 2) physical rehabilitation exercise (intervention). Within each block the search words were combined with OR. In a third block, randomized controlled trials were identified by combining two specific search strings with OR. The search strings used were Cochrane Highly Sensitive Search Strategy for identifying randomized trials (1) supplemented with a search filter based on McKibbon et al. (2). A combination of the two search strings was chosen to reach as high sensitivity as possible. Subsequently the results within the blocks were combined with AND.
As recommended, to improve quality of the search and minimize risk of bias the search strategy was peer reviewed by an experienced healthcare librarian (3), who also performed the searches in the main databases (MEDLINE, Embase, CINAHL and CENTRAL).
To exemplify the process, the MEDLINE search history from January 2024 is shown below.

#### Search History Medline THA (23.1.2024)

Search with AND | Search with OR | Delete Searches | Refresh Search Results

| Search ID# | Search Terms | Search Options | Actions |
|---|---|---|---|
| S68 | S21 AND S48 AND S66 | **Limiters** - Publication Date: 20200901-<br>**Search modes** - Boolean/Phrase | View Results (1,669)<br>View Details<br>Edit |
| S67 | S21 AND S48 AND S66 | **Search modes** - Boolean/Phrase | View Results (5,821)<br>View Details<br>Edit |
| S66 | S49 OR S65 | **Search modes** - Boolean/Phrase | View Results (7,687,711)<br>View Details<br>Edit |
| S65 | S64 NOT S63 | **Search modes** - Boolean/Phrase | View Results (4,756,347)<br>View Details<br>Edit |
| S64 | S57 OR S62 | **Search modes** - Boolean/Phrase | View Results (4,961,332)<br>View Details<br>Edit |
| S63 | PT case report OR letter OR historical article OR review of reported cases OR multicase review | **Search modes** - Boolean/Phrase | View Results (1,601,063)<br>View Details<br>Edit |

| | | | | |
|---|---|---|---|---|
| ☐ | S62 | S58 OR S59 OR S60 OR S61 | **Search modes** - Boolean/Phrase | View Results (4,886,239)<br>View Details<br>Edit |
| ☐ | S61 | double blind OR double blinded OR double masked OR triple blind OR triple blinded OR triple masked OR double-blinded OR random allocation OR random allocations OR random allocated OR randomly allocated OR clinical trial OR clinical trials OR placebo* OR random* OR trial* | **Search modes** - Boolean/Phrase | View Results (2,518,317)<br>View Details<br>Edit |
| ☐ | S60 | single blind OR single blinded OR single masked | **Search modes** - Boolean/Phrase | View Results (44,091)<br>View Details<br>Edit |
| ☐ | S59 | MW drug therapy | **Search modes** - Boolean/Phrase | View Results (2,718,641)<br>View Details<br>Edit |
| ☐ | S58 | PT randomized controlled trial OR practice guideline OR clinical trial OR controlled clinical trial | **Search modes** - Boolean/Phrase | View Results (984,378)<br>View Details<br>Edit |
| ☐ | S57 | S50 OR S51 OR S52 OR S53 OR S54 OR S55 OR S56 | **Search modes** - Boolean/Phrase | View Results (745,450)<br>View Details<br>Edit |
| ☐ | S56 | (MH "Randomized Controlled Trials as Topic") | **Search modes** - Boolean/Phrase | View Results (165,973)<br>View Details<br>Edit |
| ☐ | S55 | (MH "Random Allocation") | **Search modes** - Boolean/Phrase | View Results (106,958)<br>View Details<br>Edit |
| ☐ | S54 | (MH "Placebos") | **Search modes** - Boolean/Phrase | View Results (35,925)<br>View Details<br>Edit |
| ☐ | S53 | (MH "Research Design") | **Search modes** - Boolean/Phrase | View Results (126,296)<br>View Details<br>Edit |
| ☐ | S52 | (MH "Single-Blind Method") | **Search modes** - Boolean/Phrase | View Results (33,143)<br>View Details<br>Edit |

| | | | | |
|---|---|---|---|---|
| ☐ | S51 | (MH "Double-Blind Method") | **Search modes** - Boolean/Phrase | View Results (177,110)<br>View Details<br>Edit |
| ☐ | S50 | (MH "Clinical Trials as Topic") | **Search modes** - Boolean/Phrase | View Results (201,546)<br>View Details<br>Edit |
| ☐ | S49 | ((PT randomized controlled trial OR PT controlled clinical trial OR randomized OR placebo OR MW drug therapy OR AB randomly OR AB trial OR AB groups) NOT ((MH "Animals+") NOT (MH "Humans"))) | **Search modes** - Boolean/Phrase | View Results (6,674,039)<br>View Details<br>Edit |
| ☐ | S48 | S22 OR S23 OR S24 OR S25 OR S26 OR S27 OR S28 OR S29 OR S30 OR S31 OR S32 OR S33 OR S34 OR S35 OR S36 OR S37 OR S38 OR S39 OR S40 OR S41 OR S42 OR S43 OR S44 OR S45 OR S46 OR S47 | **Search modes** - Boolean/Phrase | View Results (2,549,900)<br>View Details<br>Edit |
| ☐ | S47 | bicycl* OR cycling OR spinning OR crosstrain* OR cross-train* OR exercise bike OR walk' OR rowing OR rower OR run or runn* OR jog OR jogg* OR sport* | **Search modes** - Boolean/Phrase | View Results (711,899)<br>View Details<br>Edit |
| ☐ | S46 | (aerobic OR anaerobic OR watergym* or water-gym* OR wateraerobic* OR water-aerobic* OR gym OR gymnastic* OR hydrotherap* OR water OR aqua*) N2 (exerc* OR train* OR rehab*) | **Search modes** - Boolean/Phrase | View Results (24,637)<br>View Details<br>Edit |
| ☐ | S45 | (weight N2 (lift* OR train* or exercis*)) | **Search modes** - Boolean/Phrase | View Results (14,885)<br>View Details<br>Edit |
| ☐ | S44 | (muscl* OR neuromuscul* OR neuro-muscul* OR resistance OR endurance OR physical OR balance OR gait OR treadmill OR aquatic) N3 (train* OR retrain* OR re-train* OR exercise* OR strength* OR fitness OR condition OR activit*) | **Search modes** - Boolean/Phrase | View Results (441,525)<br>View Details<br>Edit |
| ☐ | S43 | (strength' OR exercise* OR movement* OR physical therap* OR physiotherap* OR rehabilitati*) N3 (training OR intervention OR technique* OR program* OR session* OR class* OR isometric* OR isotonic* OR isokinetic*) | **Search modes** - Boolean/Phrase | View Results (165,975)<br>View Details<br>Edit |
| ☐ | S42 | physical N1 (education or program) | **Search modes** - Boolean/Phrase | View Results (75,078)<br>View Details<br>Edit |
| ☐ | S41 | exercis* OR train* OR physical therap* OR physiotherap* OR rehabilitati* | **Search modes** - Boolean/Phrase | View Results (1,875,877)<br>View Details<br>Edit |

Is early initiated physical rehabilitation exercise superior to no physical rehabilitation exercise following total hip arthroplasty?
A systematic review and narrative synthesis

| | | | |
|---|---|---|---|
| S40 | (MH "Swimming") | **Search modes** - Boolean/Phrase | View Results (20,165)<br>View Details<br>Edit |
| S39 | (MH "Hydrotherapy") | **Search modes** - Boolean/Phrase | View Results (2,640)<br>View Details<br>Edit |
| S38 | (MH "Walking") | **Search modes** - Boolean/Phrase | View Results (42,321)<br>View Details<br>Edit |
| S37 | (MH "Bicycling") | **Search modes** - Boolean/Phrase | View Results (13,015)<br>View Details<br>Edit |
| S36 | (MH "Physical Endurance") | **Search modes** - Boolean/Phrase | View Results (20,624)<br>View Details<br>Edit |
| S35 | (MH "Weight Lifting") | **Search modes** - Boolean/Phrase | View Results (5,122)<br>View Details<br>Edit |
| S34 | (MH "Resistance Training") | **Search modes** - Boolean/Phrase | View Results (12,641)<br>View Details<br>Edit |
| S33 | (MH "Muscle Strength") | **Search modes** - Boolean/Phrase | View Results (26,880)<br>View Details<br>Edit |
| S32 | (MH "Sports") | **Search modes** - Boolean/Phrase | View Results (35,516)<br>View Details<br>Edit |
| S31 | (MH "Physical Exertion") | **Search modes** - Boolean/Phrase | View Results (57,554)<br>View Details<br>Edit |
| S30 | (MH "Physical Fitness") | **Search modes** - Boolean/Phrase | View Results (30,016)<br>View Details<br>Edit |

| | | | | |
|---|---|---|---|---|
| ☐ | S29 | (MH "Cardiorespiratory Fitness") | **Search modes** - Boolean/Phrase | View Results (3,284)<br>View Details<br>Edit |
| ☐ | S28 | (MH "Exercise Movement Techniques") | **Search modes** - Boolean/Phrase | View Results (879)<br>View Details<br>Edit |
| ☐ | S27 | (MH "Exercise Therapy") | **Search modes** - Boolean/Phrase | View Results (50,175)<br>View Details<br>Edit |
| ☐ | S26 | (MH "Exercise") | **Search modes** - Boolean/Phrase | View Results (145,364)<br>View Details<br>Edit |
| ☐ | S25 | (MH "Rehabilitation") | **Search modes** - Boolean/Phrase | View Results (18,703)<br>View Details<br>Edit |
| ☐ | S24 | (MH "Physical Education and Training") | **Search modes** - Boolean/Phrase | View Results (14,245)<br>View Details<br>Edit |
| ☐ | S23 | (MH "Physical Therapy Modalities") | **Search modes** - Boolean/Phrase | View Results (41,262)<br>View Details<br>Edit |
| ☐ | S22 | (MH "Physical Therapy Specialty") | **Search modes** - Boolean/Phrase | View Results (2,982)<br>View Details<br>Edit |
| ☐ | S21 | S2 OR S3 OR S9 OR S10 OR S11 OR S12 OR S19 OR S20 | **Search modes** - Boolean/Phrase | View Results (116,570)<br>View Details<br>Edit |
| ☐ | S20 | joint N2 (arthoplast* OR replac* OR prosthe* OR implant' OR alloplast*) | **Search modes** - Boolean/Phrase | View Results (29,456)<br>View Details<br>Edit |
| ☐ | S19 | S16 AND S18 | **Search modes** - Boolean/Phrase | View Results (70,293)<br>View Details<br>Edit |

**Appendix A**

Is early initiated physical rehabilitation exercise superior to no physical rehabilitation exercise following total hip arthroplasty?
A systematic review and narrative synthesis

| | | | | |
|---|---|---|---|---|
| ☐ | S18 | arthoplast* OR replac* OR prosthe* OR implant' OR alloplast* | **Search modes** - Boolean/Phrase | View Results (1,020,766)<br>View Details<br>Edit |
| ☐ | S17 | S13 AND S16 | **Search modes** - Boolean/Phrase | View Results (12,124)<br>View Details<br>Edit |
| ☐ | S16 | S14 OR S15 | **Search modes** - Boolean/Phrase | View Results (203,313)<br>View Details<br>Edit |
| ☐ | S15 | hip | **Search modes** - Boolean/Phrase | View Results (203,313)<br>View Details<br>Edit |
| ☐ | S14 | (MH "Hip") | **Search modes** - Boolean/Phrase | View Results (12,620)<br>View Details<br>Edit |
| ☐ | S13 | S1 OR S4 OR S5 OR S6 OR S7 OR S8 | **Search modes** - Boolean/Phrase | View Results (31,519)<br>View Details<br>Edit |
| ☐ | S12 | THA | **Search modes** - Boolean/Phrase | View Results (17,187)<br>View Details<br>Edit |
| ☐ | S11 | THR | **Search modes** - Boolean/Phrase | View Results (23,222)<br>View Details<br>Edit |
| ☐ | S10 | hip replacement | **Search modes** - Boolean/Phrase | View Results (14,607)<br>View Details<br>Edit |
| ☐ | S9 | hip arthroplasty | **Search modes** - Boolean/Phrase | View Results (31,957)<br>View Details<br>Edit |
| ☐ | S8 | TJA | **Search modes** - Boolean/Phrase | View Results (2,002)<br>View Details<br>Edit |

**Appendix A**

Is early initiated physical rehabilitation exercise superior to no physical rehabilitation exercise following total hip arthroplasty?
A systematic review and narrative synthesis

| | | | | |
|---|---|---|---|---|
| ☐ | S7 | TJR | **Search modes** - Boolean/Phrase | View Results (587)<br>View Details<br>Edit |
| ☐ | S6 | joint replacement | **Search modes** - Boolean/Phrase | View Results (12,695)<br>View Details<br>Edit |
| ☐ | S5 | joint arthroplasty | **Search modes** - Boolean/Phrase | View Results (7,255)<br>View Details<br>Edit |
| ☐ | S4 | (MH "Joint Prosthesis") | **Search modes** - Boolean/Phrase | View Results (10,766)<br>View Details<br>Edit |
| ☐ | S3 | (MH "Hip Prosthesis") | **Search modes** - Boolean/Phrase | View Results (25,867)<br>View Details<br>Edit |
| ☐ | S2 | (MH "Arthroplasty, Replacement, Hip") | **Search modes** - Boolean/Phrase | View Results (35,410)<br>View Details<br>Edit |
| ☐ | S1 | (MH "Arthroplasty, Replacement") | **Search modes** - Boolean/Phrase | View Results (6,717)<br>View Details<br>Edit |


1. Lefebvre CG, J.; Briscoe, S.; Littlewood, A.; Marshall, C.; Metzendorf, M-I.; Noel-Storr, A:, Rader, T.; Shokraneh, F.; Thomas, J.; Wieland, L.S. Technical Supplement to Chapter 4: Searching for and selecting studies. In: Higgins, JPT; Thomas, J; Chandler, J; Cumpston, MS; Li, T; Page, MJ; Welch, VA (editors) Cochrane Handbook for Systematic Reviews of Interventions Version 6 (updated July, 2019) [Internet]. Cochrane, 2019. Available from: www.training.cochrane.org/handbook.
2. McKibbon KA, Wilczynski NL, Haynes RB. Retrieving randomized controlled trials from medline: a comparison of 38 published search filters. Health Info Libr J. 2009;26(3):187-202.
3. Lefebvre CG, J.; Briscoe, S.; Littlewood, A.; Marshall, C.; Metzendorf, M-I.; Noel-Storr, A:, Rader, T.; Shokraneh, F.; Thomas, J.; Wieland, L.S. . Chapter 4. Searching for and selecting studies. In: Higgins, JPT; Thomas, J; Chandler, J; Cumpston, M; Li, T; Page, MJ; Welch, VA (editors) Cochrane Handbook for Systematic Reviews of Interventions version 60 (updated July 2019) [Internet]. Cochrane, 2019. Available from: www.training.cochrane.org/handbook.
