## Appendix B for "Is early initiated physical rehabilitation exercise superior to no physical rehabilitation exercise following total hip arthroplasty? A systematic review and narrative synthesis"

### Supplementary results

**Table A.** Reasons for excluding studies that based on previous systematic reviews might appear eligible

| Study | Reason for exclusion |
|---|---|
| Johnsson, 1988 (1)<br>Heiberg, 2012 (2)<br>Beck, 2019 (3) | Wrong intervention (PRE was performed between discharge and study start) |
| Bodén and Adolphson 2004 (4) | Wrong intervention* (early versus late weight-bearing) |
| Morishima, 2014 (5) | Wrong intervention (initiated at a later phase after surgery) |
| Liebs, 2010 (6) | Wrong comparator (control group performed PRE) |

* Also, both groups performed home-based PRE

**Table B.** CERT checklist for studies included in the study entitled: "Is early initiated physical rehabilitation exercise superior to no physical rehabilitation exercise following total hip arthroplasty? A systematic review"

| Section/Topic | Checklist item | Study | | |
|---|---|---|---|---|
| | | **Monaghan et al.(7)** | **Martinez et al.(8)** | **Marcu et al.(9)** |
| WHAT: materials | Detailed description of the type of exercise equipment (e.g. weights, exercise equipment such as machines, treadmill, bicycle ergometer etc) | A step bench, two chairs, dumbbells, 2.5 kg weights and resistance (no further details) were used. | RSPG<br>Atabilometric Platform (AccuGaitTM 50 Hz SP (AMTITM), Watertown, MA, USA) with Balance TrainerTM (version 1.4.1) software (AMTITM, Watertown, MA, USA<br><br>SDHRG<br>Chair, couch, stair/step, and a wall | No information |
| WHO: provider | Detailed description of the qualifications, teaching/supervising expertise, and/or training undertaken by the exercise instructor | Training was provided prior to commencement of classes in the form of a practical workshop, and written illustrated manuals were provided | RSPG<br>Provided by a physiotherapist. No further description<br>SDHRG: First session supervised by a physiotherapist, otherwise | No information |

| | | | | |
|---|---|---|---|---|
| | | that included an exercise log book. | self-directed. No further descriptions | |
| HOW: delivery | Describe whether exercises are performed individually or in a group<br>Exercises are performed in classes at three community hospital-based clinical sites | Exercises are performed in classes at three community hospital-based clinical sites | RSPG<br>Exercises on the Stabilometric Platform was performed individually<br><br>SDHRG<br>Exercises was performed individually at home | No information |
| | Describe whether exercises are supervised or unsupervised and how they are delivered | Supervised by an experienced physiotherapist. The physiotherapist taught the patients 12 exercises and monitored form and exercise intensity, progressing the exercises as necessary. | RSPG<br>Supervised by a physiotherapist<br><br>SDHRG<br>First session supervised by a physiotherapist, otherwise self-directed | No information |
| | Detailed description of how adherence to exercise is measured and reported | Exercise log book completed by the treating therapist at each attendance | RSPG<br>No information<br><br>SDHRG<br>Telephone follow-up with a questionnaire was carried out twice a week to assess program compliance. No further description | No information |
| | Detailed description of motivation strategies | No description. However, willingness to attend classes and participate in an exercise programme are part of inclusion criteria | RSPG<br>No information<br><br>SDHRG<br>No information. | No information |

| | | | | |
|---|---|---|---|---|
| | | | However, telephone follow-up was performed twice a week to assess program compliance | |
| | Detailed description of the decision rule(s) for determining exercise progression | No description of rules. Exercises were progressed as necessary | RSPG<br>Same as below<br><br>SDHRG<br>Same as below | No information |
| | Detailed description of how the exercise program was progressed | Depending on type of exercise, progression was made by increasing either repetitions, speed, time duration, load or by supplemental moving of other body parts. | RSPG<br>The exercise regimen increased in difficulty and included static holds, mediolateral and anteroposterior movements and multidirectional target tracking tasks. All exercises were performed with visual biofeedback. Each patient had a tailored rest period between each exercise.<br>A specific protocol exists for each of the nine sessions.<br>The difficulty gradually increased in terms of:<br>- From bipedal to more unipedal work<br>- Decreasing size of targets<br>- Decreasing stability level<br>- Decreasing speed of moving targets<br>– More moving targets | No information |

| | | | | |
|---|---|---|---|---|
| | | | - Decreasing size of the COP cursor<br><br>SDHRG<br>Exercise regimen and progression was described in an exercise booklet as followed:<br>Go to the next exercise if you find it too challenging or painful.<br>Move on to an exercise labeled "progression" when the exercise becomes too easy | |
| | Detailed description of each exercise to enable replication (e.g. photographs, illustrations , video etc) | Written descriptions publicly available. Written illustrated manuals were provided for the therapists | RSPG<br>Written description available online in the paper's supplementary data (8)<br><br>SDHRG<br>Followed instructions and an explanatory photograph accompany each exercise.<br>Written description available online | No information |
| | Detailed description of any home program component (e.g. other exercises, stretching etc) | No additional exercises | RSPG<br>No additional exercises<br><br>SDHRG<br>Recommend to walk 20 to 30 minutes per day | No information |
| | Describe whether there are any non-exercise components | No information | RSPG<br>No information | No information |

| | | | | |
|---|---|---|---|---|
| | (e.g. education, cognitive behavioural therapy, massage etc) | | SDHRG<br>No information | |
| | Describe the type and number of adverse events that occurred during exercise | | RSPG<br>N=1 excluded due to pain. Not labelled as adverse event<br><br>SDHRG<br>No information | No information |
| WHERE: location | Describe the setting in which the exercises are performed | Exercise classes at three community hospital-based clinical sites. | RSPG<br>No information<br><br>SDHRG<br>Home-based exercises | |
| WHEN, HOW MUCH: dosage | Detailed description of the exercise intervention including, but not limited to, number of exercise repetitions/sets/sessions, session duration, intervention/program duration etc. | 12 exercises performed twice a week for 6 weeks. Each class session was 35 minutes in length. Dosage of each exercise is described in the paper's Supplementary data (7) | RSPG<br>Each session comprised 13-14 exercises, each with a duration between 10 and 120 seconds. Without including rest periods, the exercise sessions had a duration of 15-17 minutes<br><br>SDHRG<br>Exercises program performed once a day (6 days a week)<br>Each program comprised 8-10 exercises, which should be performed 3X10 per session with both legs. The first week, all exercises were without weight-bearing. Weight- | Daily exercises during 2 weeks |

| | | | | |
|---|---|---|---|---|
| | | | bearing exercises were introduced in week 2 and single leg stance in week 3.<br>Patients recommended to walk 20-30 minutes per day and to keep a record of other physical activities. | |
| TAILORING: what, how | Describe whether the exercises are generic (one size fits all) or tailored whether tailored to the individual | The exercises are generic, but the progression seems to be based on the individual participant's progress. | RSPG<br>No tailoring of exercises, but tailored rest between exercises. No further description<br><br>SDHRG<br>The following recommendations were given, providing possibility of individual adaptions - Take time to recover between each exercise - Go to the next exercise if you find it too challenging or painful. - Move on to an exercise labeled "progression" when the exercise becomes too easy | No information |
| | Detailed description of how exercises are tailored to the individual | Same as above | Same as above | No information |
| | Describe the decision rule for determining the starting level | No information | No information | No information |

| | | | | |
|---|---|---|---|---|
| | at which people commence an exercise program (such as beginner, intermediate, advanced etc.) | | | |
| HOW WELL: planned, actual | Describe how adherence or fidelity to the exercise intervention is assessed/measured | No information | RSPG:<br>Number who completed the exercise protocol. No further information.<br><br>SDHRG:<br>The first session was supervised by a physiotherapist who made sure that patients understood the instructions and distributed individual exercise booklets.<br>No specific information about fidelity.<br>Assessments with the protocol's were done with weekly telephone follow-up questionnaires | No information |
| | Describe the extent to which the intervention was delivered as planned | No information | RSPG<br>14 of 16 completed the exercise protocol.<br><br>SDHRG<br>All 11 patients completed more than 50% of the exercises that was programmed for each week | No information |

Abbreviations: RSPG: rehabilitation with a stabilometric platform group, SDHRG: self-directed home-based rehabilitation group,

**Appendix B**

Is early initiated physical rehabilitation exercise superior to no physical rehabilitation exercise following total hip arthroplasty?
A systematic review and narrative synthesis

**Table C**. Trial characteristics and results of the randomised controlled trials awaiting classification.

| **Publica-tion status** | **Trial**<br>Author, Year<br>Country<br>Study design<br>Trial registration and identifier | **Participants (intervention/ control)**<br>Numbers<br>Age (years)<br>Gender<br>Other* | **Intervention**<br>Initiating point<br>Duration<br>Short description** | **Control**<br>Short description** | **Outcomes**<br>Reported outcomes distributed on applicable domains*** | **Follow-up**<br>Time points | **Main results**<br>(as reported by the authors) | **Unclarified questions – reasons for not yet being classified** |
|---|---|---|---|---|---|---|---|---|
| Published paper | Jin et al.(10)<br><br>2023<br><br>China<br><br>RCT | N=156/156<br><br>Age in mean (SD)<br>44.6(14.2)/42.8 (13.5)<br><br>Gender proportions of females:<br>68(44%)/76(49 %) | Continuous nursing: guidance for the recovery of muscle function, psychological support, emotional support and pain control.<br>Duration: 3 months | Routine nursing | *Patient-reported function*<br>HHS<br><br>*Pain*<br>NRS | 3 months after surgery | *Patient-reported function*<br>HHS at 3 months, mean (SD):<br><br>Interventions group:<br>73.98 (10.83)<br>Control group:<br>58.77 (13.6)<br>Between group difference:<br>p=.000<br><br>*Pain*<br>NRS at 3 months mean (SD):<br><br>Interventions group:<br>3.83 (2.56)<br>Control group:<br>3.21 (1.77)<br>Between group difference:<br>p=.015 | 1) How many patients received THA due to osteoarthritis?<br><br>2) Were the patients in the control group prescribed or instructed in any specific exercises or activities? |

**Appendix B**

Is early initiated physical rehabilitation exercise superior to no physical rehabilitation exercise following total hip arthroplasty?
A systematic review and narrative synthesis

| | | | | | | | | |
|---|---|---|---|---|---|---|---|---|
| Published abstract | Matei et al. (11)<br><br>Romania<br><br>2016 | No: 16/15 | Physiotherapy exercise group (other terms used: kinetotherapy group, early rehabilitation patients group)<br><br>Based on the aim, content of the intervention is kinetic exercises and massage. | Standard care group | *Performance based function:* Muscle strength<br><br>*Quality of life:* SF-36 | No information | Compared to the usual care group, the patients with kinetotherapy program showed a better correlation between the clinical and functional variables, with a superior predictability for ROM and muscle strength in connection to pain (R = 0.712 and R2 = 0.545); moreover, the early rehabilitation patients group showed a signififcant correlation with almost 60 % (58.6) predictability for improving the quality of life. | 1) Is it a randomized controlled trial?<br><br>2) Is exercise the main intervention?<br><br>3) Was the study initiated within 3 months after surgery?<br><br>4) Were the patients prescribed or instructed to perform exercises in the period between discharge and study start?<br><br>5) Were the patients in the standard care group prescribed or instructed to perform any specific exercises or activities? |

**Appendix B**

Is early initiated physical rehabilitation exercise superior to no physical rehabilitation exercise following total hip arthroplasty? A systematic review and narrative synthesis

| | | | | | | | | |
|---|---|---|---|---|---|---|---|---|
| Pre-registered trial | Wu (12)<br>China<br><br>Parallel interventional study<br><br>ChiCTR2100041695 – (registered 2021) | Planned sample size n=60<br><br>Age ≥60 years | Rehabilitation exercise based on Cognitive Load Theory | Conventional care | The hip function scale<br>(not specified if it is a patient-reported or performance-based function outcome) | First day after surgery, at discharge and 1 month after surgery | No results | 1) Were the patients randomly assigned to the control group and the experimental group?<br><br>2) How many patients received THA due to osteoarthritis?<br><br>3) Did the experimental group receive the intervention during admission or after discharge (or both)? If after discharge: Was the study initiated within 3 months after surgery? If after discharge and |

| | | | | | | | | |
|---|---|---|---|---|---|---|---|---|
| | | | | | | | | study initiated within 3 months: Were the patients instructed to perform exercises in the period between discharge and study start?<br><br>4) Were the patients in the control group, receiving conventional care, instructed to perform any specific exercises or activities? |

Abbreviations: SD: Standard deviation; RCT: Randomized controlled trial; HHS: Harris Hip Score; NRS: Numeric Rating Scale Scores; ROM: Range of motion; SF-36: 36-Item Short Form Survey
